# Differences in medication reconciliation interventions between six hospitals: a mixed method study

**DOI:** 10.1101/2021.11.26.21266902

**Authors:** C C M Stuijt, B J F vd Bemt, V E Boerlage, M J A Janssen, K Taxis, F Karapinar

## Abstract

**Background:** Although medication reconciliation (MedRec) is mandated and effective in decreasing preventable medication errors during transition of care, hospitals implement MedRec differently.

**Objective:** Quantitatively compare the number and type of MedRec interventions between hospitals upon admission and discharge, followed by a qualitative analysis on potential reasons for these differences.

**Methods:** This explanatory retrospective mixed method study included patients from six hospitals and various wards in case MedRec was performed both on hospital admission and discharge. Information on pharmacy interventions to resolve unintended discrepancies and medication optimizations were collected. Based on these quantitative results, interviews and a focus group was performed to give insight in MedRec processes. Descriptive analysis was used for the quantitative-, content analysis for the qualitative part.

**Results:** On admission, patients with at least one discrepancy varied from 36-95% (mean per patient 2.2 (SD± 2.4) Upon discharge, these numbers ranged from 5-28% while optimizations reached 2% (admission) to 95% (discharge).The main themes explaining differences in numbers of interventions were patient-mix, healthcare professionals involved, location and moment of the interview plus embedding and extent of medication optimization.

**Conclusions:** Hospitals differed greatly in the number of interventions performed during MedRec. A combination of patient-mix, healthcare professionals involved, location and timing of the interview plus embedding and extent of medication optimization resulted in the highest yield of MedRec interventions on unintended medication discrepancies and optimizations. This study supports to give direction to optimize MedRec processes in hospitals.

## Introduction

Medication errors upon hospital admission and discharge are common and can lead to preventable adverse drug events (ADEs). ^1,2^ To diminish these errors, medication reconciliation(MedRec) is recommended in many countries: several studies have demonstrated substantial error reductions in medication errors, specifically medication discrepancies, and, to some extent, in ADEs. ^3,4,5,6,7^

In fact, MedRec consists of three different steps which are described by the Healthcare Improvement Institute (USA): 1. verification (eliminating discrepancies between a patients’ actual medication use and in-hospital prescriptions by comparing medication overviews and interviewing patients), 2. clarification (medication optimization, e.g. start of a laxative if an opioid is prescribed, eliminating double medications) and 3. reconciliation (discussion of step 1 and 2 and reasons for medication changes with the physician and documentation)(Appendix box 1a).^6^

Despite this clear definition, there is remarkable diversity in the reported effect of MedRec. For example, in the verification step detected discrepancies between the actual medication use of a patient and the medication list constituted in the hospital, was found to vary between 3.4 and 98%. ^8,9,10,11^ This broad range may be explained by variances in study methodology, differences in study population (eg acute admissions, elderly and high numbers of admission medication) but also in dissimilarities in definition. ^12^ The latter may result in different interpretations or implementation of the distinct steps of MedRec.^11,12,13,14^ E.g. the clarification step is not frequently implemented: it may be included in verification- and reconciliation steps without explicit reporting or not at all been executed^15,16^. Knowledge on inter-hospital variability of MedRec processes upon hospital admission and discharge, may give insight in MedRec-strategies and the impact on numbers of interventions. This will give direction to optimize MedRec processes in hospitals.^11,12^

Existing literature describes the impact of available staff, the employee who performs MedRec, hospital stay duration and number of admission medications on the number of interventions. However, no study performed a broader, in depth analysis to compare MedRec processes between hospitals in a real-world setting. ^13^ Hence, the objective of this mixed-method study is to quantitatively compare the number and type of MedRec interventions between hospitals upon admission and discharge, followed by a qualitative analysis of potential reasons for these differences.

## Methods

### Study design

This explanatory mixed method study consisted of a quantitative and a qualitative part.

### Ethics

The Nijmegen ethics committee reviewed the study and confirmed compliance with the Dutch legislation by giving the waiver of approval (registration nr 2013/328)

## QUANTITATIVE PART

### Setting

A retrospective cohort study was performed. Selected hospitals executed MedRec for at least five years, both upon hospital admission and discharge. As this study focussed on MedRec in a real-world setting, included wards varied based on the MedRec activities of each hospital (orthopaedics, surgery, pulmonary diseases, internal medicine and cardiology; table 1).

**Table 1.** hospital and overall included patient characteristics.

| *Hospital* | **A** | **B** | **C** | **D** | **E** | **F** |
| --- | --- | --- | --- | --- | --- | --- |
| *County (region)* | Capital (West) | Drenthe (North) | Gelderland (East) | Zuid Holland (West) | Utrecht (Centre) | Limburg (south) |
| *surrounding* | Urban | Rural | Rural | Urban | Urban | Urban |
| *Type* | Teaching | General | Specialized | General | Teaching | Teaching |
| *Number of beds 2012* | 551 | 284 | 317 | 722 | 1102 | 536 |
| *Wards with MR activities by pharmacies in 2012* | Lung diseases Internal medicine Cardiology Neurology | All wards including ICU, paediatrics | All wards | 75% all admissions covered; discharge counselling if patient passes by outpatient pharmacy or on request physician | All wards except ICUs | All wards except ED |
| *Study wards and number of patients included (n)* | Lung diseases (150) | Internal medicines (27)<br>Cardiology (50)<br>Surgery (73) | Orthopaedics (150) | Internal medicines (23)<br>Gastroenterology (4)<br>Surgery (114)<br>Lung diseases (8) | Internal medicines (107)<br>Gastroenterology (12)<br>Surgery (28)<br>Lung diseases (3) | Internal medicines (85)<br>Lung disease (65) |
| *length of stay, median (range)* | 9.0 (3-48) | 4.0 (1-60) | 6.0 (1-115) | 6.0 (1-21) | 7.0 (2-32) | 7.0 (2-37) |
| *number admission medications, mean (SD)* | 9.1 (4.5) | 6.4 (3.4) | - ‡ | 8.5 (4.1) | 10.8 (4.3) | 7.8 (4.3) |
| *number discharge medications, mean (SD)* | 10.9 (4.8) | 8.2 (4.0) | 11.1 (4.1) | 8.8 (3.6) | 10.4 (4.3) | - |
| *Number admission/discharge interviews\** | 108/149 | 140/92 | 145/136 | 147/149 | 78/106 | 146/- |
| *age, mean (SD)\*\** | 67.2 (13.3) | 62.1 (16.5) | 61.2 (12.9) | 69.3 (12.4) | 66.1 (14.6) | 69.1 (13.0) |
| *female, n (%)\*\** | 44 (41) | 80 (57) | 97 (67) | 84 (57) | 46 (59) | 79 (54) |
| *low social class, n (%)\*\** | 81 (75) | 83 (59) | 46 (32) | 57 (39) | 13 (17) | 85 (58) |
| *deprived area, n (%)\*\** | 48 (44) | 0 | 2 (1) | 10 (7) | 4 (5) | 4 (3) |

age, number of admission- and discharge medications, length of stay showed significant differences between hospitals (p=0.000)
‡ no differentiation on admission or pre-admission medication was possible
\* Admission: 134 patients had no interview, 69 (51%) patients of which were admitted to hospital E and met their exclusion criteria for medication reconciliation, 44 (25%) were incapable of being interviewed without the presence of a caregiver or had a language barrier and 21 (16%) of which had the MedRec interview upon being discharged due to a short length of stay.
Upon discharge, 632 (70%) patients were interviewed and included. Patients from hospital F (n=150, 16%) were excluded due to incomplete documentation regarding intervention performance, 117 patients (15%) were missed due to an unexpected discharge.
\*\* on admission, discharge figures were comparable and changed in a non-significant manner

### Study population

All consecutive patients admitted to a selected hospital were included if a patient had medication intended for chronic use before admission. Patients received MedRec both upon hospital admission and discharge. All included patients had had at least a discharge interview, those without an admission interview (eg due to a short length of stay) were included in case the interview could be executed upon discharge. Patients incapable to be interviewed e.g. with a language barrier were excluded. Also, patients living in an institutionalised setting before admission were excluded (presuming dependence in medication administration with consequent inability of assessing a medication history from the patient or their proxy). Per hospital we included 150 patients (900 total) based on a previous analysis on the same data.^11^

### Medication Reconciliation

At the time of inclusion most patients (>90%) used one community pharmacy (CP)^17^. Here, prescriptions from multiple prescribers are documented. Therefore, a medication history from the CP combined with a patient interview is considered gold standard to obtain the Best Possible Medication History (BPMH) in the Netherlands^18,19^. In case of doubt the GP could be consulted. Generally, the medication history of the CP is electronically available, otherwise it was obtained by fax.^15,20^

MedRec is fulfilled as described in appendix box 1a and performed by (specialized) pharmacy technicians with background support of pharmacists. Pharmacy technicians have shown to perform MedRec accurately in the Netherlands. ^21^ They have had a three year intermediate vocational training, which involves a combination of study at a college or open learning, in addition to practical working experience. A pharmacy technician can specialise further into pharmaceutical consultants, who have received an additional 3 year bachelor training focused on pharmacotherapy and communication.

### Quantitative outcomes

Number and type of interventions in the verification - and clarification step of MedRec, i.e. resolving unintentional medication discrepancies and optimization of pharmacotherapy upon admission and discharge.

### Data collection

All data were collected by three trained data collectors from hospital patient records and admission/discharge pharmacy checklists. Participating pharmacists and pharmacy technicians documented proposed medication changes (interventions) on their checklist, and, if possible communicated with the physician in charge of the patient, who would follow or reject the advice. Accepted interventions were counted, non-accepted interventions were regarded as either an intentional medication change or a non-relevant suggestion for an intervention. In case interventions were not clearly documented, pharmacy teams were available to clarify or, if no explanation could be provided, discarded.

Each accepted discrepancy or optimization was counted as an unique intervention. A single drug could therefore induce several interventions, e.g. restarting furosemide on admission to correct a discrepancy and adjusting the dose upon discharge back to the dose used at home after a temporal dose increase. In case a discrepancy between the CP list and patient reported use of medicines was noticed (including potentially stopped medication during admission), this was checked in the electronic medical record and documented as stated by the patient with discussion remarks. In case of doubt about the correct recall on medicines use of the patient, additional measures were implemented (e.g. asking for medication boxes, contacting GP).

Hospitals defined *unintended medication discrepancies* as differences among medication regimens i.e., between actual use of a patient’s home regimen and medications prescribed upon admission or discharge.^22^

*Optimizations of medication* entail a check on whether the medication list is adequate and optimal regarding (high risk) medication (eg NSAID use in combination with gastro-protection in elderly) and on duplication of therapy, based on guidelines. Both discrepancies and optimizations were recorded as described by Karapinar et al. and classified into the categories as described in appendix box 1b: *start, dosage, switch* and *discontinuation*.^15,16^

The following covariates were collected: number of chronic drugs (based on BPMH upon hospital discharge) and demographic data including age, gender and social class (via postal-code). ^23 24^ Postal-codes were also used for deprived neighbourhoods as registered by The Dutch Healthcare Authority (NZA).^25^

### Analysis

Statistics were executed with SPSS 23. Total and mean numbers of interventions both on medication discrepancies and optimizations per patient were calculated in patients with either an admission-, or discharge interview. Mean (standard deviation) or median (range) were determined dependent on the distribution of data. To compare hospitals regarding their case-mix, normally distributed continuous variables including age, number of admission- and discharge medications was analysed by ANOVA. For analysis on differences in gender, social class, deprived area, length of hospital stay, ward and admission type, the Pearson Chi-square test was used.

## QUALITATIVE PART

### Design

A qualitative explorative study using individual interviews and a focus group interview, was conducted. For the data analysis a content analysis was performed.

### Participants

For the interviews, responsible pharmacists (1 to 2 per hospital) and pharmacy technicians (2 - 4 per hospital) from included hospitals participating in the quantitative part of the study and present at the time of investigators visit, were eligible.

Involved pharmacy team members were invited to attend the focus group discussion. Five pharmacists participated in person, one was contacted by phone.

### Data collection

The qualitative part consisted of an inventory questionnaire to get knowledge on (hospital)pharmacy components of MedRec activities and feasibility of data collection. Questions included were for instance: how was the MedRec process performed (eg were all 4 steps implemented) and what type of staff members performed MedRec (see table 2 for all topics). The questionnaire was completed during an in person, semi-structured interview with participants. All interviews were conducted by the first investigator (female, pharmacist, > 20 years’ experience in community -, outpatient- and hospital pharmacy). The questionnaire was developed based on literature, experience and expert-discussion (FK, BvB). To understand the causes for differences in numbers of MedRec interventions, an explanatory focus group was conducted. During this focus group, we reached a point of ‘theoretical saturation’ as focus group members could not provide new concepts regarding the differences in the quantitative results.

**Table 2.** categories and sub-classifications extracted from interview and focusgroup.

| WHO | A | B | C | D | E | F |
| --- | --- | --- | --- | --- | --- | --- |
| Patient selection: | Non | Non | Non | Non | High risk patients* | Exclusion UDS** |
| Type of pharmacy team members involved: | Pharmaceutical consultant | Pharmacy technician + pharmacist check | Pharmacy technician + pharmacist check<br>Nurse | Pharmacy technician + specialized pharmacy technician + pharmaceutical consultant<br>Nurse | Specialized pharmacy technician + pharmaceutical consultant + pharmacist | Pharmacy technician |
| WHERE (location interview) | A | B | C | D | E | F |
| *Admission* |  |  |  |  |  |  |
|  | On the ward | Outpatient clinic (elective) | @ home (mail/phone) | Elective patients: @ home (mail/phone) | @ home (mail/phone) | Outpatient clinic (elective) |
|  |  | On the ward | On the ward | On the ward | On the ward | On the ward |
| *Discharge* |  |  |  |  |  |  |
|  | On the ward | Outpatient pharmacy<sup>†</sup> | On the ward<br>Outpatient pharmacy | Outpatient pharmacy | On the ward<br>Outpatient pharmacy + @home by phone | Outpatient pharmacy |

| HOW (process) | A | B | C | D | E | F |
| --- | --- | --- | --- | --- | --- | --- |
| **Admission** |  |  |  |  |  |  |
| Verification:<br>Information collection | digital or paper-based pharmacy dispensing information and GP | Digital pharmacy dispensing information | Patient list, extra check high risk patients***, lab results | Digital pharmacy dispensing information | digital or paper-based pharmacy dispensing information and GP | digital or paper-based pharmacy dispensing information |
| Clarification:<br>Optimisation medication | 8 focuspoints, generally discussed @ discharge | Thrombo-prophylaxis, oral antidiabetics | see above: focus on renal function, pain meds | No extra checks | see discharge | no extra checks |
|  | Substitution |  | No substitution |  | Substitution |  |
| Suggested medication changes | implementation of medication changes after check with physician | implementation of medication changes by pharmacy without physician check | implementation of medication changes after check with physician | Only acutely based on prescriptions | discussion with doctor after medication review | Only acutely based on prescriptions |
| Documentation | On electronic dispensing information from pharmacy and checklist (not visible for other healthcare professionals) | On checklist (not visible for other healthcare professionals) | On Checklist (not visible for other healthcare professionals) | ? not visible for other healthcare professionals | In EPD and on electronic dispensing information from pharmacy (partly available for other healthcare professionals) | Electronic pre-registration (elective patients) available for other healthcare professionals |
| **Discharge** |  |  |  |  |  |  |
| Optimisation (number of | 8 | 2 | 2 | 0 | 6<sup>††</sup> | 3 (surgical ward only) |
| focuspoints) |  |  |  |  |  |  |
| **WHEN (timing activity)** | **A** | **B** | **C** | **D** | **E** | **F** |
| **Admission** |  |  |  |  |  |  |
| Interview elective patients (moment) | Day admission | Day admission | Weeks before procedure by mail | Weeks before procedure | Weeks before procedure by phone | Weeks before procedure |
| **Discharge** |  |  |  |  |  |  |
| Interview will be performed | If discharge announced 24h beforehand | Only if patient passes by outpatient pharmacy | Always | Only if patient passes by outpatient pharmacy | If discharge announced 24h beforehand | Only if patient passes by outpatient pharmacy |

\* Patient interview only in case of >3 chronic drugs + 50 years or over (70-80% of all patients)
\*\* patients with drugs dispensed in unified dosing systems (UDS, Baxter)) are not counselled
\*\*\* extra check for high risk patients right before admission (age > 65 years and > 5 chronic medications). All patients fill out their own medication lists before the elective admission
<sup>†</sup> An *outpatient pharmacy* is a pharmacy based in the outpatient clinic of a hospital with community pharmacy activities, generally with close connections with the hospital pharmacy
<sup>††</sup> all issues during medication review if applicable + check on renal function and electrolytes

To ensure correct interpretation of collected information, all interviewees were sent the outcomes of the questionnaire to provide feedback on the data (member check).

An independent moderator (KT) led the group and FK acted as an observer to document relevant contributions that would not necessarily have been picked up by audio-recording. Interviewees gave oral consent for audio recording and anonymity was guaranteed. Before the meeting, all attendees were provided with drafts of the table and figures to get a clear overview of processes in each centre, patient characteristics and numbers and types of interventions. KT proposed to touch upon the following themes (based on differences between hospitals in the quantitative results) : differences in processes, persons performing MedRec and patient-mix.

The audio-recording was transcripted verbatim by an official transcriptor. Also, a member check was conducted by sending participants a summary of the transcripts in order to provide feedback on the factual and interpretive accuracy of the data.

### Qualitative Outcome

Explanations for differences in numbers of MedRec interventions by a focus group discussion.

### Analysis

Based on the group discussion, an inductive, content analysis was applied on the transcripts. First, relevant text fragments were selected individually by three researchers (CS, FK, BB) and compared to ensure no data would be missed. Second, the first researcher performed the open coding of the fragments, and applied axial coding. Relationships between the open codes were identified with axial coding and the codes were labelled into themes. This process was reviewed in its entirety by two researchers (FK, BB) until all researchers fully agreed on the content of the themes.

## Results

### QUANTITIVE PART

Overall, 899 of 900 patients were included (one exclusion due to missing information on medication use). On admission 765 (85% of 899) and upon discharge 632 (70%) patients received MedRec (table 1).

Most participants were elderly patients with a low social class (50%) (mean age 65 years, gender equally distributed) and were admitted on internal medicines, lung diseases and surgery. Overall, acute-, and elective admissions were equally distributed. However, individual hospitals varied highly in the proportion of acute admissions (0-100%), median length of stay (4 - 9 days), mean number of admission medications (6.4 -10.8) and social class (17-75% in the lowest category) (table 1).

#### Number and types of interventions

Overall, 2309 (74%) interventions were accepted by physicians; 1675 interventions (in 765 interviewed patients) were collected on admission and 634 (632 patients) upon discharge.

##### Unintended discrepancies (verification)

On admission, the proportion of patients with at least one discrepancy varied between hospitals from 36% to 95%. Upon discharge, this ranged from 5% to 28% of patients.

Overall, hospitals varied with regard to the distribution of types of interventions. In general, start and dosage interventions were most frequently implemented, as medication a patient used pre-admission was omitted or patients used different dosages. Also, stops and switches were needed: medication was prescribed that the patient did not use anymore (commission error) or patients used another drug at home (e.g. pantoprazole instead of omeprazole).

Upon discharge, start interventions were performed most frequently in the majority of the hospitals (e.g. restart of pre-admission medication that was temporarily discontinued).

##### Optimizations of medication

On admission, the proportion of patients with at least one optimization varied from 0 to 27% and upon discharge, this ranged from 2 to 95% of patients. Highest numbers of optimizations were found in hospitals where the clarification step was integrated into the MedRec process, specifically on discontinuation of medication (e.g. discontinuing hypnotics that were initiated during admission and had no indication anymore upon discharge)(figure 2).

### QUALITATIVE PART

#### MedRec differences between hospitals

Analysis of the audio-transcript of the focus group emerged in three themes: *who* performed MedRec, *where* and *how* was MedRec performed (table 2).

*Who* (interviewer, patient mix, physician type)

> <u>interviewer</u>: all participants agreed that highly trained MedRec interviewers, resulted in high numbers of pharmacy interventions (although hospital A, with the highest numbers of interventions, had both higher educated and highly trained interviewers). Therapeutic knowledge of medicines use was judged key to apply properly optimization checklists (see below) in order to remove inappropriate and unnecessary medications from a patient’s medication list.
>
> <u>patient-mix/physician type</u>: participants agreed that surgical patients generally have less discrepancies versus general wards as they use less medication. Also, participants reflected that a high social class would result in less discrepancies due to the higher education level. This could explain the high amount of interventions in hospital A with 75% of patients having a low social class and 100% patients from the pulmonary department. In contrast: hospital B, C and D included a substantial percentage of surgical patients resulting in the lowest number of interventions upon discharge (table 1, figures 1,2). According to participating pharmacists, surgeons generally will not act upon optimization interventions.
>
> Figure 1 mean number and type of interventions per patient due to unintentional discrepancies upon admission and discharge * discharge excluded due to the high number of interventions without documentation on acceptance. adm: admission dis: discharge 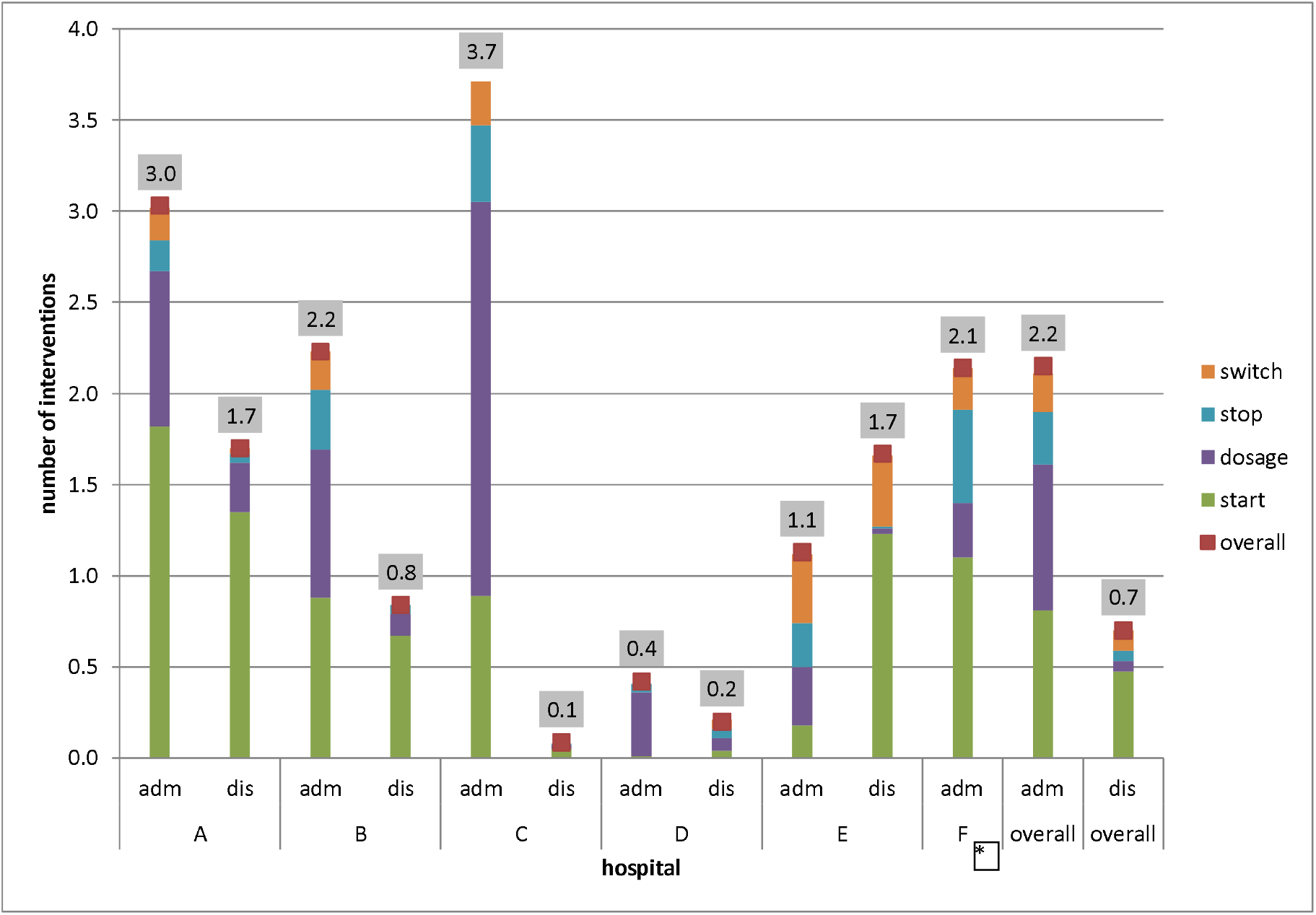
>
> Figure 2 mean number of optimizations per patient per hospital upon admission and discharge * discharge excluded due to the high number of interventions without documentation on acceptance. Adm: admission Disc: discharge 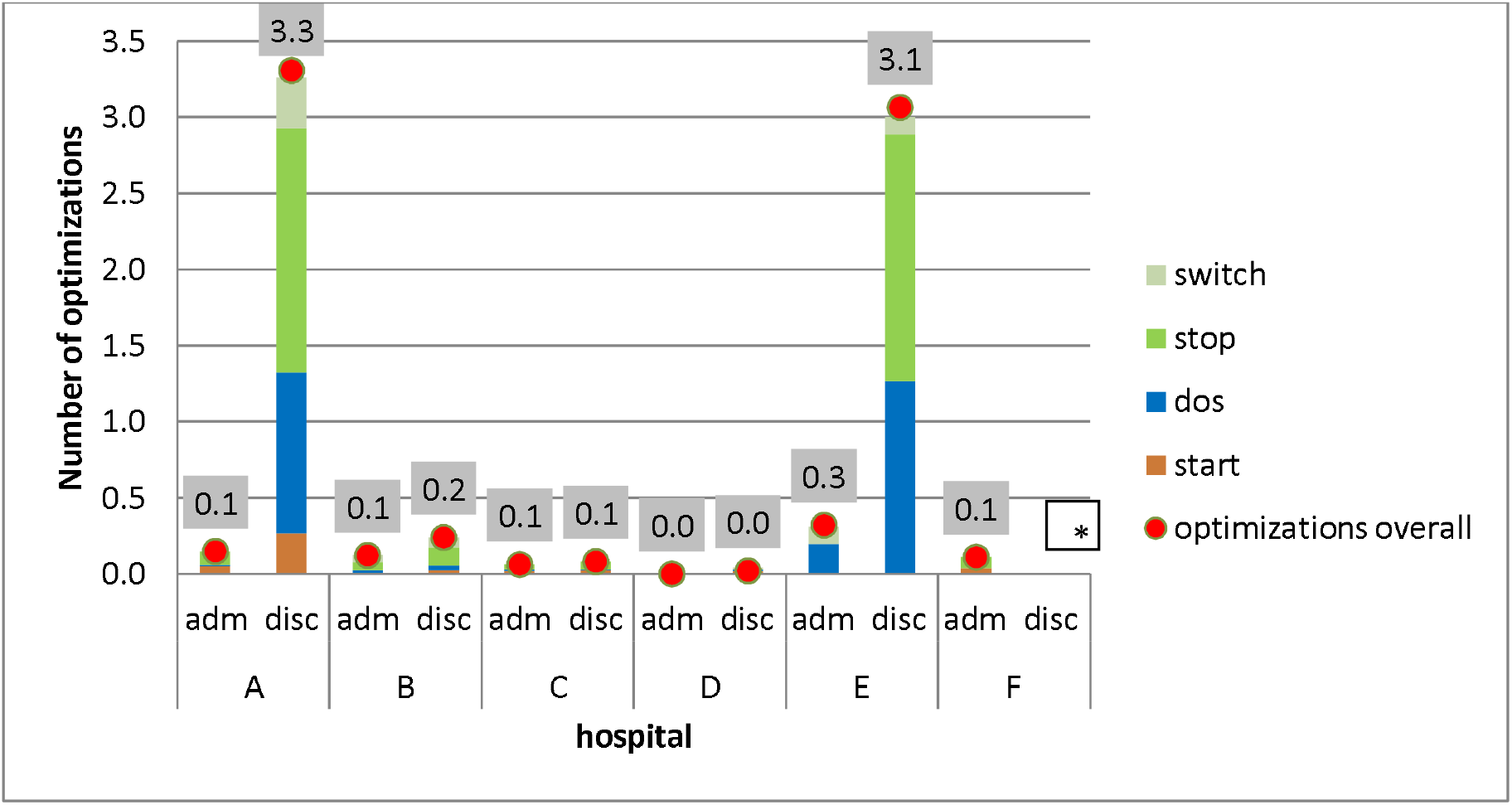
>
> Hospital A and E had very comparable workflows but had different numbers of interventions on discrepancies: 3.0 versus 1.1 on admission (figure 1). Included patients differed highly: lung diseases only (hospital A), as compared to 70% of internal medicine patients (hospital E, table 1). In participants’ opinion, internal medicine doctors pay more attention to medication, probably resulting in a lower number of discrepancies (less found by the pharmacy team), even though high numbers of medication were used.

*Where* (at home, outpatient pharmacy or on the ward)

> <u>Pre-admission preparation at home:</u> a form filled out by the patient himself (instead of using the CP medication history to perform MedRec) several weeks before elective admission, resulted in a shift in type of interventions from start to dosage (figure 1, hospital C and D). Patients appeared to recall which medicines were used, but failed to note or remember dose and/or strength of a medication. This resulted in the high number of dosage interventions when MedRec was performed as compared to omissions in all other centres.
>
> <u>Location of the interviews (ward versus outpatient pharmacy)</u>: two hospitals, B and D, discussed discharge medication and counselled patients in the outpatient pharmacy only, instead of ward-based counselling. Those hospitals had at least 50% less interventions on discrepancies as compared to hospitals with ward-based MedRec and/or telephone interviews. This might have been the result of a less intense connection with in-hospital activities, according to participants.

*How* (optimizations and documentation)

> <u>optimizations:</u> hospitals differed in numbers and types of medication optimizations (0-8 checks structurally included) eg the extensiveness of the checklist resulted in large differences in numbers of optimization interventions (figure 2). Furthermore, in hospital E, two pharmacists performed a medication review for selected patients (elderly, more medications) which, potentially contributed to the higher number of optimization interventions, specifically upon discharge. This higher yield was explained by acceptance of certain clinical situations (in the context of medication use) while being admitted and were discussed during medication review while action and documentation was postponed to discharge (e.g. accepting potassium suppletion in combination with potassium sparing medications while being admitted, but not in the outpatient setting without frequent lab control).

## Discussion

### Key findings

In this real-world setting study, quantitative and qualitative comparison of MedRec processes in 6 different hospitals revealed considerable differences in numbers of interventions: upon admission, patients with at least one discrepancy varied from 36-95%, while upon discharge, these numbers ranged from 5-28%.Optimizations reached 2% (admission) to 95% (discharge). Based on the qualitative analysis, we identified three main themes explaining differences between hospitals: patient- and healthcare professionals involved, where the interview was started or performed and to what extent medication optimization was embedded in the process.

### Comparison with previous work

In a recently published report (among 19 hospitals) the large variability between hospitals on performance of all steps was confirmed.^26^ In this report describing highly involved hospitals and pharmacy teams, the variation was explained by obstacles like resource shortage, available staff and enthusiasm of the management. Comparable results were found in a study of the MedRec process in four acute care hospitals: varying levels of compliance with guidelines were noticed e.g. interview with the patient occurred in less than half of all MedRec (45.7%).^27^ Other important barriers for MedRec implementation mentioned were: lack of awareness and insufficient knowledge of health care professionals.^13^ Apart from aforementioned obstacles and high risk patient population (elderly with or without polypharmacy), other factors like, social class and process-related factors have not been mentioned or investigated in previous studies. Therefore, our results give new insight in facilitators and barriers to perform MedRec activities within hospitals.

### Implications for practice

This retrospective cohort without robust analysis should be interpreted with caution prior to further research. However, some specific topics may have the potential of a quick win in daily practice: differences in types of interventions were noticed amongst electively admitted patients having had the ability to fill out their medication list before admission. In that case, the majority of MedRec interventions were on dosage adaptions, whereas in situations lacking this patient list, medication was mostly started due to MedRec. Hence, the most common intervention type changed from omission to dosage, which might decrease the severity of the error from moderate to mild.^28^ Furthermore, an optimization checklist increased the number of interventions on general wards. This effect on inappropriate medication use by standardization of a medication chart, especially with a high number of focus points, has been proven previously.^29,30^ However, no such effect was noticed on surgical wards in our study. This was explained by the fact that surgeons generally will not act upon optimization interventions and the high number of general, non-surgical focus points in the checklist.

Introduction of medication review (for selected patients) increased the intervention numbers as compared to the other hospitals (figure 1), giving room to a further improvement of health problems in daily live. ^31^

Upon discharge, some hospitals had very low numbers of interventions. Two potential explanations were given:1. high numbers of surgical patients in combination with 2. MedRec performed solely in the outpatient pharmacy (plus discharge medication dispensing). Probably, to prevent a surgical patient from medication related problems, either an admission interview only might be sufficient, (as reflected by the high number of discrepancy interventions on admission in hospital C) and/or a closer connection with the clinic (both pharmacy teams and physicians) may result in better communication and acceptance of more interventions.

### Strengths and limitations

To our knowledge this is the first multi-center analysis on MedRec processes providing insight into differences in interventions on medication discrepancies and - optimizations. Inclusion of a large and varied population (eg different patient categories and physician-types) increased generalizability and transferability; the connection of quantitative and qualitative results created the ability to improve our understanding of MedRec and give input for new research.

However, several limitations have to be mentioned. First, this study was performed in the Netherlands, reducing its external generalisability due to differences in health care systems. However, we expect that the main drivers between differences in MedRec interventions will not extremely differ between countries. Not all possible factors influencing the extent of MEdRec are measured in the quantitative part of this study (eg specific patient characteristics like disease burden and workload of those who perform MedRec). However, the qualitative synthesis gave important insight and augmented also new facilitators and barriers for MedRec.

Third, documentation between hospitals differed and the retrospective nature of the study gave rise to a reporting bias as interventions were not always clear. This could result in an underestimation of the total number of interventions. However, this was not very frequent except for hospital F where we excluded the discharge results. Fourth, the actual impact of prevention of medication discrepancies on the individual patient has not been analysed. Therefore, we do not know if these discrepancies would have resulted in patient harm. However, Mekonnen et al. showed that a pharmacist-led MedRec program upon hospital transitions decreased ADE-related hospital revisits, all-cause readmissions and ED visits, resulting in a positive impact of MedRec.^32^

## Conclusion

Hospitals differed greatly in the number of interventions performed during MedRec. Upon admission a variation of 0.4-3.7 interventions per patient was noted. Upon discharge the variation was 0.1-1.7 interventions per patient. A combination of patientmix, healthcare professionals involved, location and moment of the interview plus embedding and extent of medication optimization resulted in the highest yield of MedRec interventions on unintended medication discrepancies and optimizations.

## Supporting information

Appendix Boxes

## Data Availability

All data produced in the present study are available upon reasonable request to the authors

## Acknowledgements

The investigators would like to thank participating hospital pharmacists and technicians. These are: E Helfrich, Department of Pharmacy, Wilhelmina Hospital, Assen; M Meijs, Department of Pharmacy, Sint Antonius Hospital, Nieuwegein; S v Bommel and R Smit, Department of Pharmacy, Albert Schweitzer Hospital, Dordrecht; S vd Meer and S Dings, Department of Pharmacy, VieCuri Hospital, Venlo, all in the Netherlands, for their cooperation in this study. Furthermore we express gratitude to the following students for collecting data: D Bervoets, R de Rooij, N Wartenberg.

## Statements and declarations

The authors have no conflicts of interest to declare. They declare to have no financial or non-financial interests that are directly or indirectly related to the work submitted for publication.

The dataset generated during and/or analysed during the current study is available from the corresponding author on reasonable request.

