## Appendix Boxes for "Differences in medication reconciliation interventions between six hospitals: a mixed method study"

Appendix Box 1

**Box 1a: medication reconciliation process**

1. On admission:

a. Verification: collection (within 24 hours) of a (pre-admission) community pharmacy medication history that contains a 6 months history of dispensed medication which is combined with a patient (or their proxy) interview in order to create the best possible medication history (BPMH). This is compared with the list of medications ordered upon admission to the hospital. During this step the right drug, dose, route and frequency of administration is checked. In case patients are unable to be interviewed, a paper based medication reconciliation is performed comparing the medication history of the community pharmacy with the medication prescribed in the hospital.

b. Clarification*: medication and dosages are checked for appropriateness by the pharmacy technician based on guidelines (e.g add laxative if an opioid is prescribed or adjust medication based on kidney malfunction, proton pump inhibitor indicated if NSAID is introduced or already in use).

c. reconciliation: based on aforementioned information a medication list is documented by the pharmacy technician. Any of the non-ordered, inappropriate or changed medications are discussed with the physician. Hereafter, the patient's medication records are updated either electronically or paper based, to facilitate access for the physician, explicitly documenting changes and reasons therefore.

2. At discharge:

a. Verification: medication reconciliation is performed using admission medication reconciliation information (as mentioned in 1), hospital data and patient information for the comparison and detection of discrepancies with the medication prescribed at discharge.

b. Clarification: medication and dosages are checked for appropriateness by the pharmacy technician based on guidelines (e.g. is there still an indication for hypnotics, analgesics or laxatives started in the hospital)

c. Reconciliation: based on this information a discharge medication overview specifying medication changes and reasons, is created by the pharmacy technician after discussion with the physician on possible unintended discrepancies, undocumented changes in medication in discharge medication instructions and prescriptions.

d. Transfer: this discharge medication overview is provided to the next healthcare provider (e.g. general practitioner/community pharmacist). Patients are counselled regarding medication changes.

* two hospitals performed a semi –structured medication review.

Medication optimisations were performed through checking on safety and quality of pharmacotherapy eg the continuing need for a medication, identification of sub-optimal treatment and identification of clinically relevant drug-drug or drug-disease interactions.

**Box 1b classification discrepancies and optimizations**

1. start (discrepancy: incorrect deletion of a medication used before admission, optimization: guidelines advise to start a drug)
2. dosage (discrepancy: difference in dosage, scheme, strength or formulation used before admission, optimization: guidelines advise a change, e.g. in patients with decreased kidney function)
3. switch (discrepancy: unintentional change in drug in the same therapeutic group, optimization: guidelines advise another medication in the same therapeutic group
4. discontinuation (discrepancy: a drug was not used before admission, optimization: a drug does not have an indication anymore, e.g. temporarily intended medication in hospitals)
